# Characterizing parametric differences between the two waves of COVID-19 in India

**DOI:** 10.1101/2021.06.29.21259698

**Authors:** Arpit Omprakash

**Affiliations:** Department of Biological Sciences, Indian Institute of Science Education and Research Mohali, Mohali – 140306, India

## Abstract

The first case of COVID-19 in India was reported on January 30, 2020 [1]. The number of infections rose steeply and preventative measures such as lockdowns were implemented to slow down the spread of the disease. Infections peaked around mid-September the same year and the cases gradually started declining. Following the relaxation of lockdown and the appearance of mutant strains of the virus, a much severe second wave of COVID-19 emerged starting mid-February. For characterization and comparison of both the waves, a SIQR (Susceptible-Infected-Quarantined-Removed) model is used in this paper. The results indicate that a single patient can infect approximately 2.44 individuals in the population. The epidemic doubling time was calculated to be 11.8 days. It is predicted that the actual number of infected patients is grossly underestimated (by a factor of 16) by current testing methods.

## INTRODUCTION

The second wave of COVID-19 has been described as an overwhelming “tsunami” [2] that took over the nation. As of June 25, 2021, total number of cases stands at 30 million with over 350,000 deaths, highlighting the intensity of the second wave compared to the first wave in India.

Mathematical modelling has played a critical role in understanding and predicting the impact of the pandemic at global and regional scales. Compartmental models such as the SIR (Susceptible-Infected-Removed) models have been proposed to study the transmission, growth, and decline of communicable diseases in a given population. Due to the pre-emptive testing drives and mandatory quarantine of confirmed positive individuals, a variation of the SIR model - called the SIQR(Susceptible-Infected-Quarantined-Removed) model - has been proposed as a viable model to study the evolution of COVID-19 [3]. This modelling approach has been used widely to study the effects of the first wave of COVID in many countries, including Italy [4], Brazil [5], and India [6].

In this paper, analysis and characterisation of parameters for the growth and spread of the second wave of COVID-19 in India has been done.

## SIQR MODEL

The SIQR model has four compartments - Susceptible, Infected, Quarantined, Removed (*Figure 1*). Individuals in the ‘Susceptible’ (S) compartment who acquire the disease from an infected person upon contact are moved to the ‘Infected’ (I) compartment. Infected individuals may be symptomatic or asymptomatic. Symptomatic infected individuals that test as positive are placed in the ‘Quarantined’ (Q) compartment, assuming that asymptomatic infected ones do not take the test at all. Finally, individuals that recover or succumb to the disease are moved into the ‘Removed’ (R) compartment.

**Figure 1:**
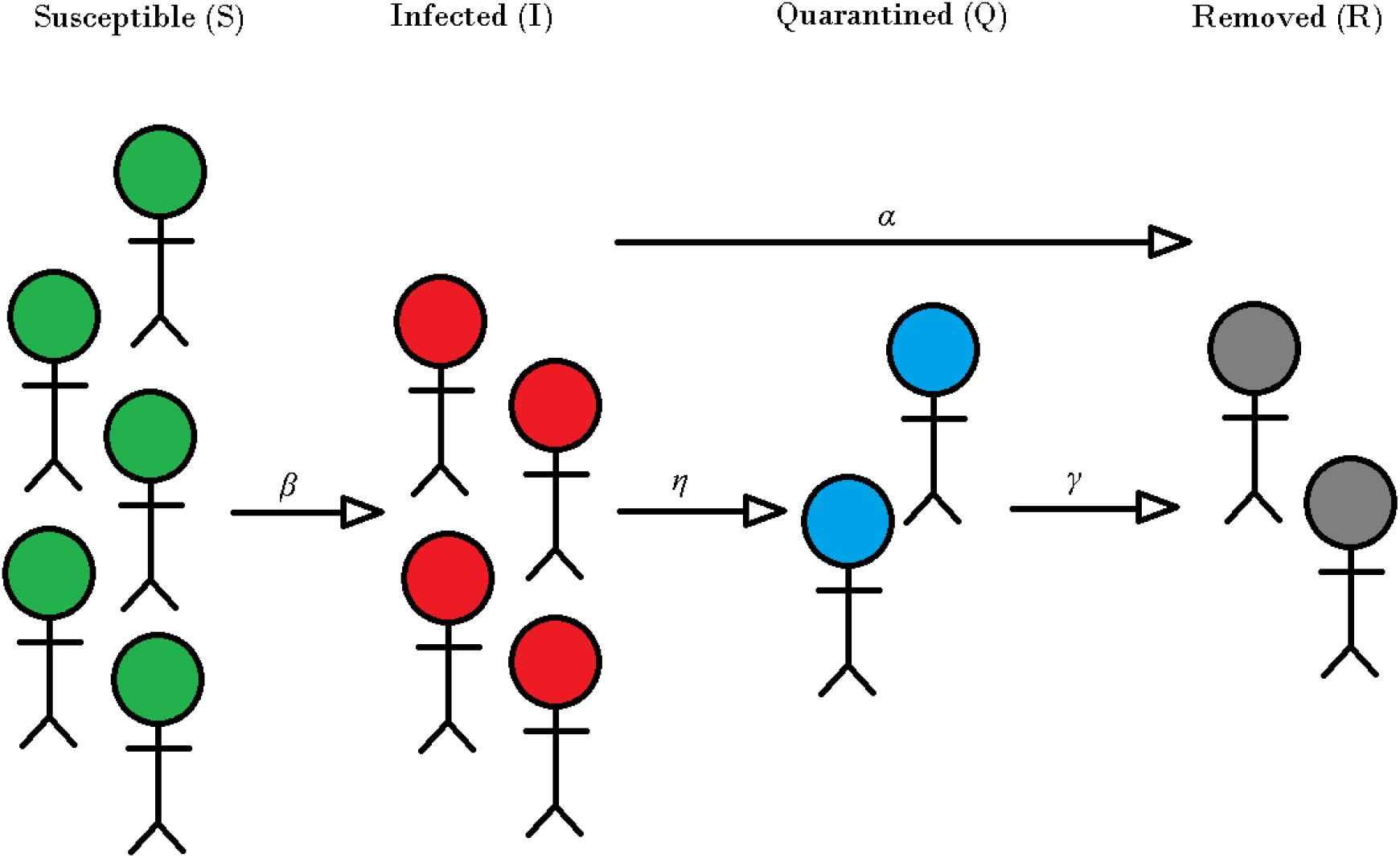
The SIQR model. The S, I, Q, and R compartments contain proportion of population that is Susceptible, Infected, Quarantined, and Removed respectively. β, η, α, and γ are rate of infection, rate of detection, rate of removal of asymptomatic cases, and rate of removal of quarantined individuals from the population respectively.

The equations describing change in states (or compartments) in the SIQR model is given below [7]:

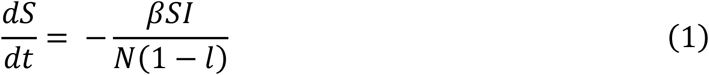

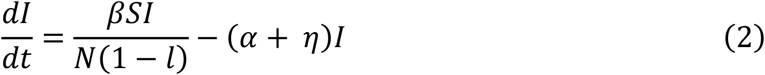

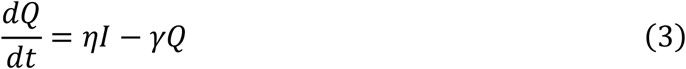

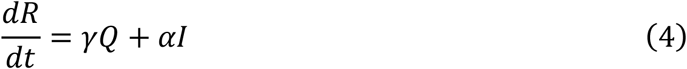

where,

*N* is the population size

*β* is the rate of infection

*η* is the rate at which individuals who tested as positive are quarantined

*α* is the rate of removal of infected people who were not quarantined

(asymptomatic or untested symptomatic)

*γ* is the rate of removal of quarantined people from the population

*l* is the fraction of population following lockdown protocol

## ANALYSIS and RESULTS

Modified equations of the SIQR model are fitted with the reported data of the country [8].Assuming that the population not following lockdown is susceptible, i.e., *S ≈ N*(1 − *l), Eq. 2* can be approximated to

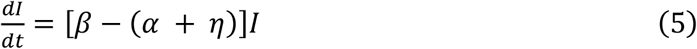

Integrating Eq. 5, we obtain

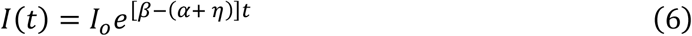

Since the beginning of the second wave was reportedly around mid-February, day 0 for the analysis is attributed to February 13, 2021 [9]. As the wave finally dies down with declining number of cases towards the first week of May, the fourth of May is considered as the last day for the analysis (t = 80).

Substituting t = 0 in *Eq. 6, I*(*t*) i.e., number of infected people at t = 0 is Io. Calculating the number of infected people on February 13, 2021, as *number of confirmed cases – (number of individuals cured + number of deaths), I*_*o*_ = 1,36,571 [8].

Adding *Eq. 3* and *Eq. 4*, the rate of change of total number of confirmed cases in the country is given by:

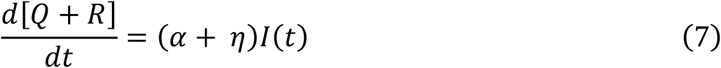

Substituting the value of *I*(*t*) from Eq. 6

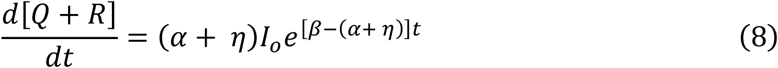

Integrating, we obtain

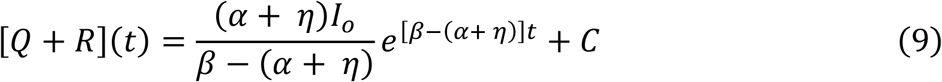

*Eq. 9* is of the form 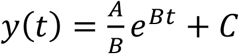 where *y*(*t*) is the number of confirmed positive cases on day ‘t’. This form of the equation is fitted to the data using least square fitting (*Figure 2*) to give *A* = 5503.0, *B* = 0.0583, and *C* = 10827627.9. Using *I*_*o*_ = 136571 and calculating values for the parameters, (*α* + *η*) = 0.0403, and *β* = 0.0986.

**Figure 2:**
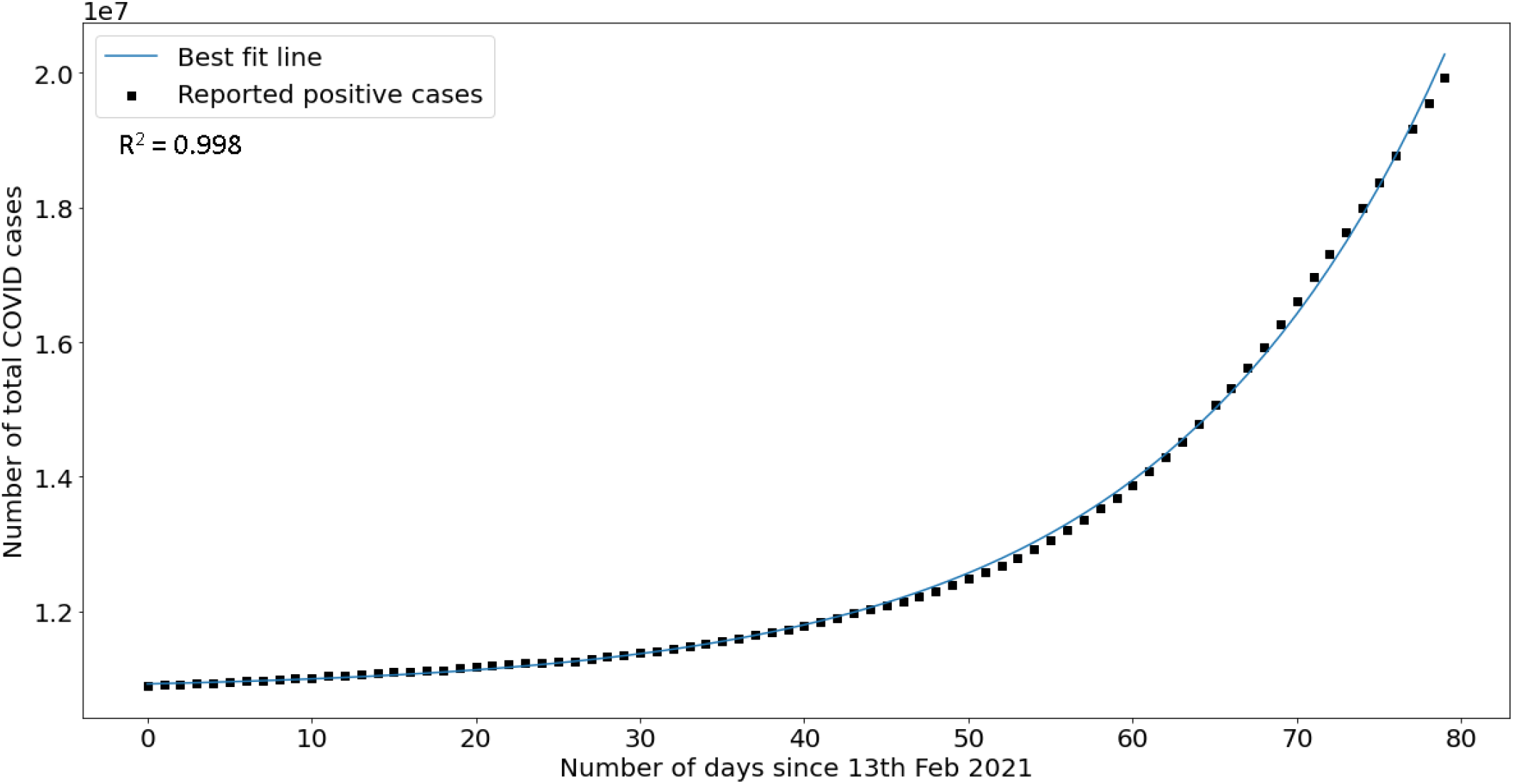
Least square fitting of Eq. 9 with total confirmed positive cases from February 13 – May 4, 2021.

Assuming that all the individuals that tested positive were quarantined, the number of active positive cases on a given day ‘t’ is given by *Q*(*t*). Substituting the value of *I* from *Eq. 6* in *Eq. 3*, we get

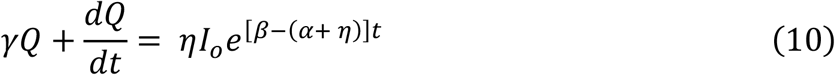

Integrating *Eq. 10* over time,

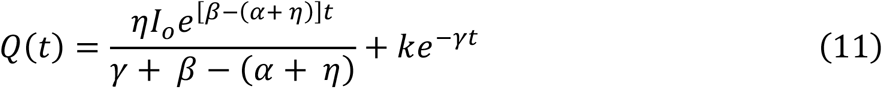

*Eq. 11* is of the form 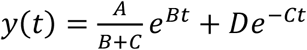 where *y*(*t*) is the number of confirmed active cases on day ‘t’. Using least square fitting, this form of the equation is fitted to the actual data (Figure 3) to give *A* = 4029.95, *B* = 0.05, *C* = 0.43, and *D* = 4917.79. Calculating the values of the parameters, *η* = 0.0295, *α* = 0.0108, and *γ* = 0.43.

**Figure 3:**
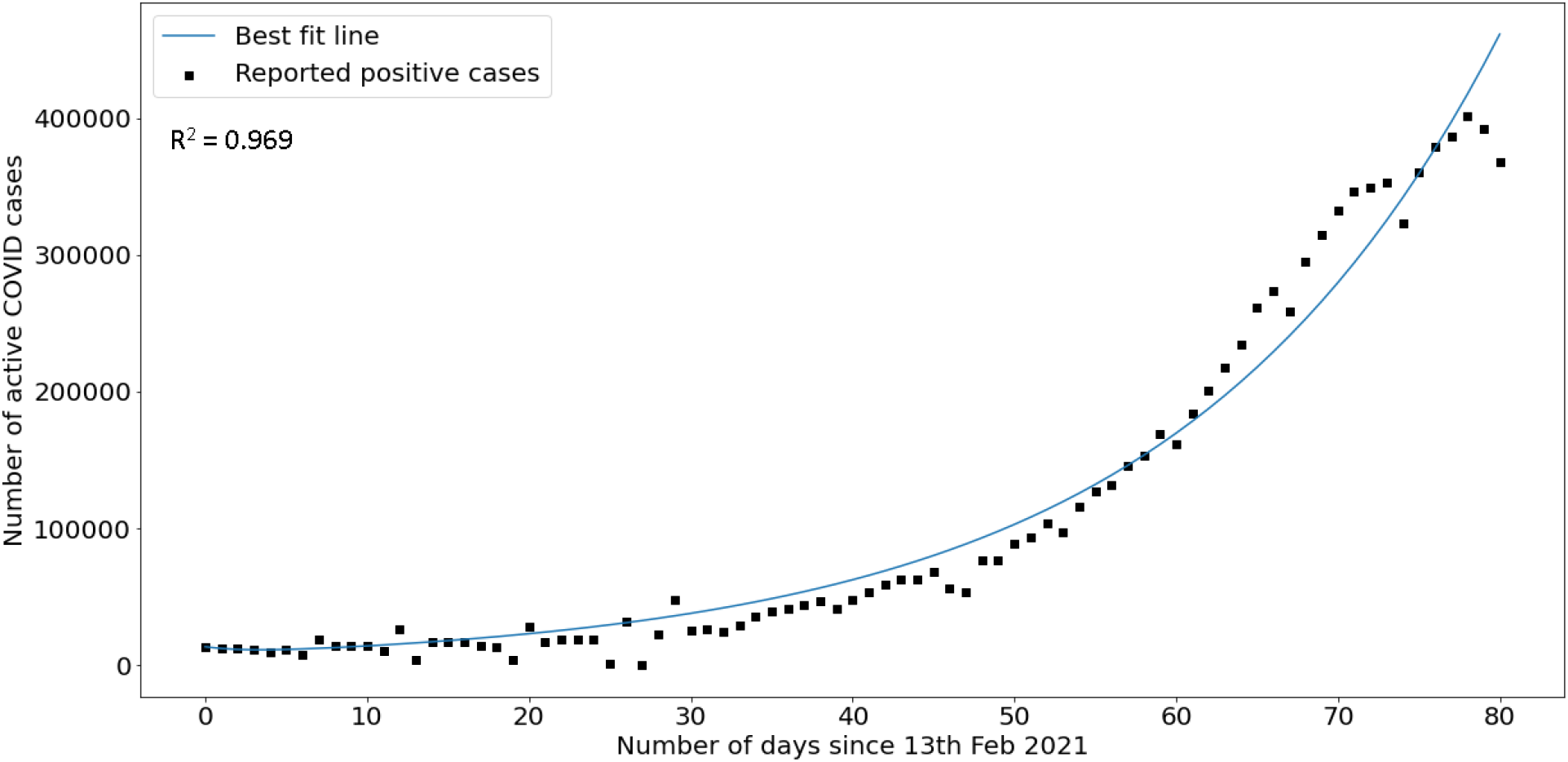
Least square fitting of Eq. 11 with active positive cases from February 13 – May 4, 2021.

The estimated parameters can be used to determine indicators that characterise the transmissibility of the disease [6]. The following indicators were calculated:

- Reproduction Number (*R*_*o*_) – The reproduction number is defined as the average number of individuals that a single infected individual can infect. It is calculated as:

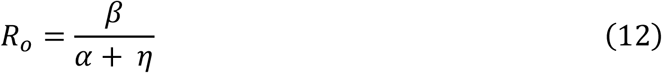
- Epidemic Doubling Time (*τ*) – The epidemic doubling time is the amount of time (in days) required for a disease to double the infected population. It is calculated as:

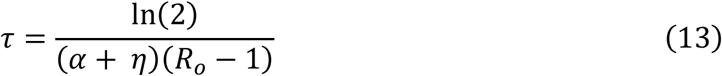
- Infected to Quarantine ratio – Ratio of the actual number of infected individuals in a population (symptomatic and asymptomatic) to the ones that are quarantined. This ratio is given as:

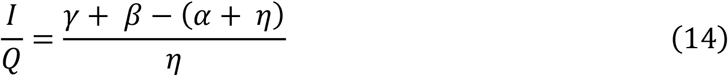

The values for these indicators (along with values for the first wave in India determined here [6]) are outlined in the table below:

## DISCUSSION

The value for *β* − (*α* + *η*) is estimated as 0.0583 in the first curve fitting, and 0.05 in the second curve fitting. Both values are close and any minor difference can be attributed to the difference in the goodness of fit (for the former R^2^ = 0.998 while for the latter R^2^ = 0.969), indicating that the approximations concur. The epidemic doubling time has increased when compared to the first wave which may be due to the larger load of cases in the second wave compared to the first wave, implementation of lockdown measures, vaccinations, and following of covid appropriate behaviours by individuals.

The values for the reproduction number (*Table 1*) indicate that the second wave of covid is more infectious compared to the first wave. The estimation agrees with the emergence of highly contagious strains such as B.1.617 [10], [11] in India. Also, increased exposure of younger individuals [12] to the virus has led to a rise in the number of susceptible individuals and possible infections. The infection to quarantined ratio for the second wave is times higher compared to the first wave indicating the presence of a greater number of carriers for the disease (asymptomatic/mildly symptomatic individuals who have not been tested or quarantined). The ratio is indicative of the low testing per million in India [13], where the highest number of tests per million is around 1276 (May 4, 2021) within the timeframe of the modelling and analysis. The positive rate of detection starts off at 1.60% on February 13, 2021, and rises up till around 21.6% towards the first week of May. The high positive rate is indicative of a steeper growth of virus in the country compared to the detected confirmed cases, and the presence of a very large number of undetected cases [13].

**Table 1:** Indicators determining transmissibility of the disease for first wave [6] and second wave of COVID-19 in India.

| Indicator | First Wave | Second Wave |
| --- | --- | --- |
| Reproduction Number | 1.55 | 2.44 |
| Epidemic Doubling Time | 4.10 days | 11.8 days |
| Infected to Quarantine Ratio | 10.45 | 16.55 |

## CONCLUSION

The SIQR model has been used to estimate the parameters and characterise parametric differences between the first and second waves of covid in India. The following points summarizes the findings of the paper:

- On an average, a single individual can infect around 2.44 other individuals (*R*_*o*_ = 2.44) which is higher than the *R*_*o*_value for the first wave in India.
- It takes about 11.8 days to double the number of cases of covid in India while it took only 4.10 days during the first wave.
- The ratio of actually infected individuals to the number of confirmed infected individuals is 16.5 which is 1.5 times higher than that of the first wave indicating the presence of a large number of undetected cases of infection.

## Data Availability

All data used in the manuscript is available online. Links to the datasets are provided along with the manuscript.

https://www.kaggle.com/sudalairajkumar/covid19-in-india

https://ourworldindata.org/coronavirus-testing

## DISCLAIMER

I am not an epidemiologist, the analysis and following discussion about the second wave is based on elementary mathematical modelling of COVID-19. The paper should not be consulted as a guiding document to take any administrative decisions.

## ACKNOWLEDGEMENTS

I wish to extend my gratitude to Subhashree Subhadarsini for proofreading and language editing of the manuscript.

